# Development of health and safety training for Vietnamese American nail salon owners and workers

**DOI:** 10.1101/2021.10.25.21265480

**Authors:** Tran B Huynh, Duong T Nguyen, Nga Vu, Catherine Freeland

**Author notes:** **Correspondence:** Tran Huynh, MPH, PhD, CIH, Dornsife School of Public Health, Nesbitt Hall, 3215 Market Street, Suite 613, Philadelphia, PA 19104.

## Abstract

**Background:** Nail salon workers are an underserved worker population that faces multiple barriers to accessing occupational health training and services. We developed a series of occupational health training modules, which were culturally tailored to Vietnamese-speaking workers, covering topics on infection control, musculoskeletal disorder prevention, chemical safety, and labor practices. We delivered the training online (due to COVID-19) to a small group of Vietnamese owners and workers in the Philadelphia metro area to obtain feedback on the training content and potential implementation challenges.

**Methods:** Seven participants (three owners and four workers) were recruited to attend the training. Qualitative feedback was obtained after each training session, followed by a more in-depth interviewer-assisted open-ended questionnaire to gain better understanding of the potential challenges of implementing the recommended changes. The Health Belief Model was used to guide the analysis of the participants’ responses to identify the perceived benefits and barriers of the training.

**Results:** Themes of perceived benefits of the training were bridging the gap of cosmetology school training, offering practical tips to protect their health at work, and inspiring conversations about work dignity and labor practices. Themes of perceived barriers were availability of affordable safer products and lack of resources, desire to please customers, lack of commitment from owners, and ubiquitous low wage that impacts employee’s job satisfaction and motivation to change, and difficulty in obtaining a work license.

**Conclusions:** Our study revealed the multitude of social and economic barriers facing immigrant nail salon owners and workers. Potential policies and strategies to overcome some of these structural barriers are discussed for the long-term health protection of nail salon workers.

## Introduction

The modern nail industry in the United States (USA) largely employs immigrant and minority workers who often earn low wages enabling consumer prices to be affordable for the masses. A report by the UCLA Labor Center and the California Healthy Nail Salon Collaborative (2018) reported that approximately 78% of nail salon employees (excluding the self-employed) earn less than $13.46 an hour and the median wage for full-time nail salon workers is around $9.06 an hour ^1^. In addition, manicurists across the country have frequently reported acute adverse health effects associated with the nail salon environment such as respiratory, eye and throat irritations, headaches, skin problems, and musculoskeletal pains (e.g., Boston, Massachusetts^2^, Alameda County, California ^3^, East Coast ^4^, Philadelphia metropolitan area ^5^, and Michigan ^6^). Prior literature has also documented potential long-term health effects such as adverse birth outcomes ^7^ and cancer^8^ among nail salon care workers, however these studies are limited and inconclusive possibly due to limited demographic information available, long latency of the diseases, and the change in the industry workforce composition ^8^. A more recent study of reproductive health effects among nail salon technicians reported strong association for congenital heart defects and neural tube defects ^9^. Additionally, manicurists are also at risk for infectious diseases such as fungal infections and bloodborne pathogens (e.g., hepatitis B and HIV) from accidental cuts and contaminated equipment.

Immigrant manicurists face significant health disparities in addition to the environmental exposures faced in the workplace. Language barriers and limited resources prevent workers and owners from accessing high quality multilingual health and safety training and occupational health services. Asians make up most of the nail salon workforce (76%), followed by white workers (14%), Black, Latinx, and other ethnicities (10%). Vietnamese specifically, dominate the nail industry while other Asian communities such as Chinese, Korean, Indian, Filipinx, and Nepali also contribute labor to the industry ^1^. The racial and ethnic diversity of workers in this industry adds another level of challenge for outreach because these workers face language and cultural barriers, lack of familiarity with the public health and health care infrastructure and labor practices.

Over the past fifteen years, awareness of poor health outcomes within the nail industry have led to increased outreach and policy implementation in some states and local governments. For example, in New York in 2015, the governor passed a comprehensive package of legislation and emergency regulations requiring nail salon owners to implement strict workplace safety standards to correct and prevent unlawful practices and unsafe working conditions ^10^. Additionally, California, adopted a voluntary approach to incentivize nail salon owners to adopt safer practices recognition as healthy nail salons within their local jurisdictions ^11^. King County, Washington ^12^ and Boston, Massachusetts ^13^ also implemented similar programs to the California safer nail salon initiatives.

Despite the pervasiveness of the issue, evidence-based intervention studies for this worker population are scarce and the use or adaptation of tested interventions for outreach is unknown. Quach et al., 2018 led the only large-scale randomized intervention study in California that used a train-the-trainer approach to work with Vietnamese nail salons to reduce chemical exposure^14,15^. Within this program owners were invited to participate in a day-long training which encouraged them to then train their workers on program lessons. The study found significant improvement in knowledge, self-reported safety behaviors and some reduction in personal exposure to harmful chemicals (total volatile organic compounds) ^14, 15^. In another study conducted by a county health department, salons were provided educational pamphlets on infection control practices and visited by outreach workers to discuss material content. The study reported a statistically significant decrease in infractions in both the intervention and control groups, suggesting the study suffered a Hawthorne effect making it difficult to determine whether the effect was due to the intervention materials or being observed by the state health inspectors visits ^16^. Another study developed and delivered health and safety training to English-speaking cosmetology trainees and found improved self-reported knowledge and work practices after 90-day post-test ^17^. With the exception of the California study, most of existing intervention studies did not address the unique cultural and linguistic needs of immigrant nail salon care workers and owners.

In order to address the limited resources and identify best practices related to environmental health prevention efforts, this study seeks to describe the development of a comprehensive health and safety training module targeting Vietnamese nail salon care staff and owners, and to report the perceived benefits and barriers to implementing recommended changes.

## Methods

### Recruitment

Vietnamese nail salon owners and workers were recruited using convenience sampling approach through a community partner network. Participants had to be at least 18 years old, working part-time or full-time at nail salons in the Greater Philadelphia area (counties located in the Southeastern Pennsylvania, South Jersey, and Delaware) at the time of the study. Potential participants were reached by phone and explained the purpose of the study. Since the study took place during the coronavirus pandemic (COVID-19) in the fall of 2020, the training was delivered remotely using the Zoom platform. During recruitment, our community partner asked participants about their technology needs and level of comfort with using Zoom. All participants had either a laptop, an iPad or a tablet and access to the internet either at home or at work. Additional in-person technology training was scheduled with our community partner on an as needed basis for five out of seven participants unfamiliar with Zoom prior to the training.

### Development of training modules

To determine training content, health and safety resources for nail salon workers from governmental sources (OSHA, EPA, FDA), relevant articles and tips from the industry trade magazine (cite), and training materials shared by the California Healthy Nail Salon Collaborative (cite) were reviewed. The final training content included four main topics: infection control, musculoskeletal diseases (MSD) prevention, chemical safety, and labor practices (Table 1). Presentations were developed in PowerPoint in English and then translated into Vietnamese. Multiple iterations of the training were co-developed with community partner staff member (a former nail salon worker) to ensure the content and translations were culturally and linguistically appropriate. The three online sessions were conducted over Zoom and included a mixture of PowerPoint slides, instructional videos, and interactive discussions.

**Table 1:** Healthy Nail Salon Training Content by Topics

| Table 1: Healthy Nail Salon Training Content by Topics |  |  |  |
| --- | --- | --- | --- |
| Infection Control | Musculoskeletal Disease Prevention | Chemical Safety | Labor Practices |
| • Common infectious diseases at nail salons<br>• Methods to properly clean and disinfect tools<br>• Methods to clean and disinfect pedicure tubs<br>• How to handle blood<br>• Basic information of Hepatitis B, transmission and risks at nail salons, common myths, testing and vaccination | • Common musculoskeletal disorders in nail salon workers<br>• Adjusting working positions, postures and movements, and basic stretch exercises<br>• Adjusting the work environment<br>• | • Common chemical substances used at nail salons<br>• Chemical exposures at nail salon<br>• Health effects of chemicals<br>• Safety Data Sheets<br>• Ways to reduce chemical exposures through substitution, ventilation, and change in work practices. | • Three levels of laws and regulations: federal, state and local<br>• An overview of OSHA, IRS, Labor Department, The Board of Cosmetology and Department of Public Health<br>• Nail salon workers' rights<br>• Nail salon owner's responsibilities<br>• Worker misclassification |

### Implementation of the training

The synchronous online training was offered on Wednesday evenings during the least busy days after work so that it would not interfere with their family time or salons’ operation on the weekend. The first training session on infection control, hepatitis B, and MSD prevention and the second session on chemical safety were offered to owners and workers together. The last session on labor regulations and worker’ rights was offered to owners and workers separately so that nail technicians would be more comfortable asking questions. All trainings were administered by our study staff and trained community partner. Each training session lasted approximately one hour and was delivered in Vietnamese language. Participants were compensated for their time and presented with a certificate of completion at the end of the training.

### Qualitative data collection and analysis

After each training session, participants were asked to join 10-minute breakout sessions with our bilingual study team to provide feedback on the content and delivery. Nail care staff were in separate breakout sessions from owners to minimize the power dynamics affecting their responses. Responses were typed by study staff and translated into English for analysis. In addition to breakout sessions, a questionnaire was administered after the training date. The questionnaire consisted of opened-ended questions based on the Consolidated Framework for Implementation Research (CFIR) to gain a comprehensive understanding of the potential implementation challenges of training. The CFIR is a conceptual framework that was developed to guide systematic assessment of multilevel implementation contexts to identify factors that might influence intervention implementation and effectiveness ^18^. The framework is often used to guide rapid-cycle evaluation of the implementation of complex healthcare delivery interventions ^19^. The CFIR has 39 constructs covering five domains: inner setting, outer setting, characteristics of individuals, intervention process, intervention characteristics, and the implementation process. Our questionnaire focused on implementation readiness constructs ^20^ to ensure the collected data were relevant to our research objectives. Our community partner assisted in the data collection by helping participants complete the questionnaire over the phone. Responses were typed in Qualtrics (^21^ and then translated into English for analysis.

The qualitative breakout session feedback and text responses from the open-ended questionnaire were independently coded by two team members (TBH and DTN) in ^22^). Major themes were categorized into the Health Belief Model constructs because we were interested in understanding the perceived benefits and barriers for implementing the suggested changes from the training. Multiple meetings were held to discuss the themes and resolved differences. A third team member (CF) joined to discuss potential differences. The project was approved by Institutional Review Board (IRB) of Drexel University.

## Results

### Demographics

Seven participants (three owners and four workers) worked in salons in various locations including New Jersey (N=3), Delaware (N=2), and Pennsylvania (N=2). Four were males and three were females. The age range was between 30 and 54 years old and the mean was 40 years (demographic table is included in the supplemental materials).

### Health Belief Model

The Health Belief Model (HBM) was used to guide our qualitative interview data analysis to understand individual knowledge and perception of workplace hazards (perceived susceptibility and severity), individual understanding of the benefits of the training (perceived benefits) and the barriers to act on the training recommendations (perceived barriers), as well as individual confidence in making changes (self-efficacy) and external events that prompt behavioral change (cues to action) ^23^

#### A. Perceived susceptibility and severity

Interviews revealed an overall lack of knowledge or understanding about chemical safety, infection control, ergonomics, and labor laws among Vietnamese nail salon workers.

Before the training, some participants perceived low susceptibility regarding their risk to catching a disease at workplace. One participant said, *“I am very relieved knowing that I went to work for all those years without getting infected by any disease. I am shocked at how easily transmitted some diseases are. I am very thankful for the training and will spend extra time cleaning between clients*.*”*

Many nail salon workers and owners who were interviewed also did not have a clear understanding of the chemical components in their nail products as well as their toxicity and potential health impact. They reported not being adequately trained in this topic during their licensing process. One salon owner, who is responsible for purchasing nail products, said: *“I have never paid attention to chemical content of the products I bought. I just bought the cheapest products available*.*”*

Moreover, many Vietnamese nail salon workers were foreign-born immigrants and not familiar with American labor laws. The majority of the participants reported not having a clear understanding of overtime pay, paid sick leaves, minimum wage, and tax laws. One worker participant said after the training on the labor laws he still could not apply much immediately because he needed to *“do more research on paid sick leaves,”* and *“if it is too complicated and [his] owners are busy, they will not do it*.*”*

#### B. Perceived Benefits of the training

Three main benefits of the training were identified from the analysis.

1. The training helps fill the gaps in current cosmetology training and provide continuous education for nail professionals. The notable gaps addressed by training includes both the inadequacies in training content specific to health outcomes and lack of training material or content in the workers’ native language (Vietnamese). *“I find [this training] very helpful. I had lessons from school in the past, but they were not this detailed and practical*.*”*
2. The training raised awareness of workplace hazards and provided nail salon care workers and owners with practical tools and techniques to protect themselves. They said the training helped them better understand different chemicals, how to choose safer products as well as how to properly handle, label and store chemicals. One participant expressed: *“[The training] provided me more awareness about chemicals and ways to protect my health and make sure everyone else is safe*.*”* They also learned about practical sanitation methods to prevent the spread of infectious diseases and were encouraged to adopt new habits to prevent musculoskeletal disorders. One participant said, “*I will be more mindful with my body position at work and remember to take breaks between customers. It might be difficult to break old habits, but I will pay more attention and put more effort into fixing it*.*”* Moreover, the training also provided practical tools to effectively communicate with customers about safety and professionally handle common situations at work, which was appreciated by most participants. One said, *“I find this very helpful. This is information that I can apply to my everyday work… I feel more comfortable and confident in dealing with situations at work… I now know how to react better, especially when my customers have a cut*.*”*
3. The training has sparked new conversations among workers and owners about workplace safety and the dignity of work. These conversations have allowed workers and owners to reimagine a new kind of nail salon where both the workers and the business can thrive. The workers in the interviews expressed their hope to raise their dignity to a new level and a sense of solidarity in helping each other. The following are some of the important questions that were raised by the workers after the training:

*“Can you do something so that the nail salon workers would be valued more?”*

*“If you are on vacation, will you have the salary or not? Right now, we don’t*.*”*

*“We need a support group in case something happens. How can we support workers to get their license?”*

For the owners, they said after the training, they would look more into paid sick leaves, overtime pay and talk with their accountants about worker classifications and taxes.

### Perceived barriers of the training

Six common perceived barriers related to the implementation of the training were identified and are described below with corresponding quotes.

1. Availability of safer products and equipment and lack of resources: Most participants believed healthy products cost more. One owner said, “*I think the costs for healthy nail salons might drive up compared with those that were not “healthy*.*”* They also expressed concerns about the cost of new equipment and materials such as new ventilation systems, local exhausts, and personal protective equipment (PPE) as well as concerns about the willingness to pay for these additional expenses. One participant said: *“When it comes to the steamer or vacuum cleaner, some salons do not even buy them in order to save money, let alone these [additional expenses]*. Some participants expressed concerns about the availability of safer products at their local nail supplies, “*I am not sure if they sell the safer products that you recommended at the nail supply I go to. Sometime the workers just don’t have any choices but to buy toxic products just because they are the ones available. In some states or locations, it’s just so hard to have access to nail supply that the choices are just limited*.” When asked about the type of support they might need to make their salons healthy, owners particularly expressed financial support “to pay for their workers”, “health insurance for workers”, “renovate their salons”, “help with 401k for workers”. Other support included “support in skills and business strategies and classes on safety and labor laws”,
2. The desire to please customers may compromise safety protocols: One participant suggested that some changes recommended in the training could be difficult to implement in practice because nail salons desire to please customers and talking about infection control for example can put people off. Another technician talked about how many practices such as cutting cuticles might not be hygienic but that’s what the customers wanted, “*The content of the training is ok, but not exactly realistic, 80-90% difficult to apply. Like when you said when we cut them, we should not touch their blood, but I feel that we need to wipe the blood for them and do it safely. I don’t want to upset the customer*.*”* In terms of the recommended cleaning protocol, one worker said, “*It’s difficult to apply cleaning suggestion. If the salon is not busy, we can clean according to the instructions you all provided. However, when the salon is busy, we have to be fast and clean much faster*” regarding the 10-minute soak in disinfectants of pedicure spa. This suggested the need for training to include more efficient cleaning suggestions such as the use of disposable pedicure liners to accommodate for busy days.
3. The need for the owners to be on board to make any meaningful difference: Most workers expressed they did not have much power to implement changes in their salons because they were not the owner. They agreed that most of their coworkers followed the instructions from the owners and they did not want to do things against the owner’s wills. They emphasized the importance of enrolling and training the owners for any meaningful difference to be made at the salon’s level. One worker said, “*This is all dependent on the owners. They have to be responsible for buying safe products. To protect my own health, I can spend my own money to buy safer product for me to use when I work with customers but sometimes the owners just don’t like it and don’t allow it to happen*.” Another worker expressed: “*Workers like me want to keep to the cleaning habit, however, the owners will also need to understand and allow time for us to thoroughly clean our tools and working stations to keep us and customers safe*.” The owner participants suggested the training needed to consider the challenges in applying changes into practice and be more realistic about the expected outcomes. One owner said, “*I think the training is appropriate for some nail salons but may be inconvenient for some others. This can be time-consuming for the owners to talk with their workers and organize the training*.” Another owner was concerned if they became a healthy nail salon, they would have to follow more regulations and standards, which could be burdensome, “*I am concerned that if we apply more requirements, the workers might not be comfortable following more rules and quit*”.
4. Low wage affects employee’s job satisfaction, which can then impact motivation for change. One worker said: “*I think it’s complicated at nail salon because most work extra hours like 9-10 hours per day for most workers. They need to work that much to earn a little bit of extra money*.” Another worker contributed: “*It’s difficult to pay overtime, sick leaves, and vacations. Both workers and owners don’t want to pay more when they already struggling*.”
5. Difficulty obtaining a license Most participants expressed not receiving much help on obtaining a license. Their businesses are also mostly family businesses. The pressure of earning a livelihood as new immigrants and language barriers put many workers and owners in vulnerable predicaments. One worker said: “*Many workers don’t have license. We tend to hire their family members to help them earn an income, like those who were studying and working*.*”* One owner expressed the dilemma they were facing: “*We don’t have the condition to follow the rules - We know that we need the license to work but the workers need to work to earn money to live. Obtaining a license is challenging for many people*.”

### Self-Efficacy

In terms of self-efficacy, workers expressed higher confidence in changing personal behavior after the training but low confidence in influencing the environment, the salon’s policy, or coworkers’ behavior. The participants overall agreed that changes that can be applied immediately to their daily practices involve personal hygiene and ergonomics techniques such as frequent handwash, use of PPE (masks, gloves…), proper working positions to prevent injury, proper cleaning, and disinfecting techniques and stretching muscles during the breaks. One participant said, “*If it involves my habits, it is easier to change than changing the salon’s setup*.”

However, in terms of training recommendations on changing workplace environment, the workers unanimously agreed that they did not have much power and confidence in influencing the salon’s policy or their coworkers’ practices. They emphasize the need for owners to be on board to make any meaningful difference. One worker said, “*It’s important to have the owners on board with any changes because they often had more power to implement and enforce changes*.*” Another said: “I am a worker and I have to care for my livelihood first. I do everything for my livelihood. I don’t have much power as a worker to change things up*.”

The workers also suggested the need for additional requirements from State Board as a catalyst for change. One participant said: “*I think you should encourage state board to do the requirement on training and safe practices*.” Most of participants agreed that collaboration with the government, State Board or public health departments will improve the credentials of the training program and facilitate enrollment and participation. One worker said, “*I think if you are doing it, you have to do it to the end, collaborate with state board and talk with the owners. If you do the training during the weekend and state board is there, people will stay and attend the training. People have to be scared a bit to follow the rules*.*”* Some believed it was better if the training is compulsory and required by State Board.

### Training logistics/delivery

The last theme from our analysis relates the training logistics and delivery. While the evening training time appeared to work for some participants with no children or with grown children, it was particularly problematic for some participants with young children as it interfered with their family dinner time, “*the time interferes with family dinner, I suggest offering the training in the mornings*.” The other challenges were the need to ask permission to leave work early (for nail technicians) or having to miss the training to keep their business open (for the owners).

Preference for the training format was mixed. On-line was preferred by some participants, “*I prefer online because it is most convenient for my work and personal schedule*”. Another individual shared, *“I think online is good for everyone because we work late and do not have time*.” Participants preferring in-person over online format expressed that it would be easier for them to comprehend the training and they would not need to deal with technology challenges, “*during COVID, we have to do it online but after COVID, in person may be better and easier to understand. Sometimes if the internet connection is slow, it’s difficult to follow*.” Another nail technician echoed similar sentiment and added that the training should be offered at the salons and made mandatory for both owners and workers, “*training at the nail salons may be better because we will have both the workers and owners. But I am not sure if all the workers will want to be trained. I think it’s better if the training is compulsory. If it’s required by state board, the owners will for sure change more*.”

Lastly, many participants noted that having the training in simple “*mother-tongue language*” made the training much easier to understand, *“it is easy to understand in Vietnamese because it is my first language*”. The participants also acknowledged the trainers for being clear, detailed, approachable, and passionate in their instructions, “*the trainer is passionate and explains in detail*”.

## Discussion

Small nail salons, like many other small businesses, often lack the knowledge and resources to implementing trainings to assist with health and safety issues. We employed community participatory approach to co-develop in-language occupational health training with our community partners and tested the training with nail salons in Philadelphia metro area. The original intention was to deliver the training in-person, but COVID-19 forced us to test the training using online format. While there were challenges helping participants navigate Zoom technology, however, we also saw tremendous opportunities in the use of online formats (synchronous or asynchronous training format) and social media platforms to reach more nail salon workers and owners in larger geographic areas in a more cost-effective way. Within the Vietnamese nail community, there are numerous YouTube how-to channels and influencers using social media to advertise their nail products and services. Many of these channels can be easily accessed with just the phones at work or home. In addition to providing technical support to overcome technology barriers, the online space makes materials accessible through social media platforms such as YouTube, Facebook and could enhance outreach efforts to accommodate busy work and family schedules. While online training may be an efficient way to deliver information to certain workforce within the nail community, on-going interpersonal technical support after the training to help owners and workers implement the changes will be important and this is where cultural and linguistic knowledge will be critical to bridging that connection.

A caveat to note is the differences in state regulations for nail salons. While federal standards on safe workplaces enforced by the Occupational Safety and Health Administration (OSHA) are appliable to all nail salons across the country, nail salon owners are most concerned with their respective State Board of Cosmetology regulations as the State Board inspectors typically conduct more inspections than OSHA. Future trainings that cross state’s boundaries should also address their state’s specific regulations to make the training more relevant to the intended audience.

In addition to providing feedback on the training content, our participants informed us the multitude of barriers beyond the occupational health training program itself that needs to be addressed simultaneously to ensure long-term positive impact on worker health and foster economic vitality of small immigrant owned businesses.

### Implications

The limited regulation of chemicals in the cosmetic industry has flooded the consumer market with a plethora of toxic products. A few ‘safer’ alternatives are often limited in availabilities and too expensive for most consumers. This is an equity issue. Yet, manufacturers continue to profit from these toxic products without much accountability and pass on the responsibility/burden of health protection from the use of their toxic products to the consumers themselves, local governments, small business owners and workers. Thus, stricter regulations on the manufacturers would prevent toxic products to be in the market in the first place and yield the greatest impact. Recognizing that individuals with low socioeconomic status, women of color and salon workers bear the disproportionate harm from toxic products, the Safety Beauty Bill Package is an example of effective advocacy targeting the upstream determinants of worker and consumer health ^24^. Introduced in October 2021 in Congress, the bill federally bans toxic chemicals in beauty and personal care products, defends women of color and salon workers by requiring translated safety data sheets of products commonly used by these communities and providing funding for development of safer alternatives, requires disclosure of fragrance and flavor ingredients, and ensures transparency the supply chain ^25^. This legislation will significantly help to address the expressed concern relating to the cost and limited availabilities of safer products in the nail salon supply chain.

Currently few states offer comprehensive in-language occupational health training to help nail salon owners and workers (e.g., California Healthy Nail Salon Collaborative ^26^, and New York Nail Salon Workers Association ^27^. Our recommended strategy is to train and empower our community-based organizations who already have been serving the nail community for other health and social needs with occupational health knowledge and skills so that they can better reach out to the nail salon workers. Not only do trainers need speak the same language as participants but also exhibit the respect and compassion for the community members and understanding of their workspace. Our participants noted a positive feature of our training was our trainers, suggesting that our community outreach educator was a significant factor in the positive reception of the training. We urge local governments and other funding agencies concerning with worker health or immigrant health disparities to invest in these community-based organizations for occupational health outreach in immigrant communities. Successful models has been replicated in several states such as the CA Healthy Nail Salon Collaborative, Adhikaar (a non-profit working with Nepali nail salon staff in New York ^28^, and Vietlead in Philadelphia. Those community-based organizations have been powerful agents in pushing for policy change and community defense at the local and state levels.

Challenges operating small businesses were a recurring theme emerged from our conversations, suggesting that in order to get the owners’ buy-in to support occupational health training, their business’ concerns also need to be addressed. Many of the worker health concerns go unaddressed within the nail industry and stem from language barriers between owners and staff, limited understanding of the labor laws, low support and investment from the larger business community and local governments. Many small nail salons are not part of a business association or local chamber of commerce further isolating them from resources. Thus, they often do not have access to business development opportunities, mentorship, or legislative influence. There is a need for the larger business community and local government partnership to invest and support the nail industry that supports so many minorities and immigrants. Example investment could be in-language business trainings and mentorship to immigrant entrepreneurs on business management and worker health, grants and low-interest loans to upgrade their salons with safer products and better ventilation.

Lastly, challenges relating obtaining a work license continues to put many immigrant nail salon workers and owners in vulnerable predicament. In Pennsylvania, trainees are required to sit in 250 hours of learning in a cosmetology school but many reported learning more and better on the job. Allowing for an apprentice program so that new immigrants can quickly earn a living while learning on the job would reduce barriers. The New York Nail Salon Worker School^29^ has a unique apprentice program to support new manicurists regardless of immigration status by offering shorter in-language training program (26 hours) and allowing for manicurists to register as apprentices and earn fair wages until they are ready to take the exam. Such program or any efforts to reduce barriers to obtaining a work license will help to reduce worker vulnerabilities and fear of authorities.

### Limitations

Our participants were all Vietnamese nail salon owners and technicians in the Philadelphia region and thus may not be generalizable to all nail salon workers, particularly those of different ethnic and cultural backgrounds. Our sample size of six might be noted as a limitation. However, given our objective was to pilot the training delivery and obtain feedback on our training modules from Vietnamese owners and workers, our target audience for a subsequent study, we felt that our objective was achieved. In addition, through our follow-up open-ended interview, we were able to obtain rich information about perceived barriers to implementing the interventions, many of which were consistent with previous published qualitative studies about nail salon workers in other regions ^5,6,30^. These insights have informed us to identify appropriate resources to support salon owners and workers following the training and engage with relevant local stakeholders such as local government, chamber of commerce, philanthropy to invest in the community to sustain and expand the program.

## Conclusions

To our knowledge, few nail salon intervention studies document the perceived benefits and barriers of implementing recommendations to make nail salons ‘healthy.’ In order to address the limited resources and best practices related to environmental health prevention efforts, this study describes the development of a comprehensive health and safety training modules targeting Vietnamese nail salon care staff and owners, and to report the perceived benefits and barriers to implementing recommended changes. This study offers an opportunity of lessons learned and feedback for future public health programming within this community and strategies and possible best practices for creating content for the Vietnamese nail salon community. The reported benefits of the training were increased self-efficacy, filling in training gaps from cosmetology schools, and sparking new conversations about health and labor practices. The study also revealed the perceived benefits and the multitude of social and economic barriers beyond the occupational health training itself that included cost and availability of safer products, the desire to please customers, lack of commitment from owners, low prices affecting employee’s job satisfaction and motivation to change, and difficulty obtaining a work license. To overcome these perceived barriers, there needs to be 1) federal mandate targeting manufacturers to minimize the production of toxic chemicals in beauty and personal care products and investment in the development of safer alternatives so that safer products can be less expensive and more available in the nail supply chain to most consumers, 2) establishment of local policy and technical support programs that are culturally appropriate and engaging local community-based organizations to address the gaps of occupational health regulations at the state and federal levels, and 3) economic investment and business mentorship for immigrant nail salon entrepreneurs to help them see the business value of operating profitable and healthy salons.

## Data Availability

All data produced in the present work are contained in the manuscript

## Authors’ contributions

TBH conceptualized the study, led the development of the training, data collection, analysis, interpretation of the results, and manuscript preparation. TBH is responsible for all aspects of the work. DTN co-developed the training, delivered part of the training, substantially contributed to the data acquisition, analysis of the qualitative data, and manuscript preparation. NG co-developed the training, recruited participants, delivered the training, and provided critical insights into the survey design, data collection, and issues facing the nail community. CF contributed to the hepatitis B training and editing of the manuscript. All authors reviewed and approved the final manuscript.

## Acknowledgements

We are indebted to the participants for their feedback on the training and opinions about issues affecting nail care professionals and industry. We are grateful of the following individuals who helped to facilitate the training: Tracy Nguyen, Hoang Anh Nguyen, Mai Anh Nguyen, Emily Nguyen, and Trisha Le. We thank Chau Nguyen for the translation of the responses. Lastly, we thank Drs. Igor Burstyn and Amy Carroll-Scott for reviewing the initial draft of the manuscript and the anonymous reviewers for their feedback.

## Institution and Ethics approval and informed consent

This study was approved by the Institutional Review Board at Drexel University’s Office of Human Research Protection. All participants provided oral informed consent.

## Disclaimer

*None*

